# Protein Interaction Networks Define the Genetic Architecture of Preterm Birth

**DOI:** 10.1101/2020.06.05.20123232

**Authors:** Alper Uzun, Jessica Schuster, Joan Stabila, Valeria Zarate, George Tollefson, Anthony Agudelo, Prachi Kothiyal, Wendy S.W. Wong, James Padbury

**Author notes:** Equal Contributions (Alper Uzun, Jessica Schuster).

## Abstract

Rather than pathogenic variants in single genes, the likely genetic architecture of complex diseases is that subgroups of patients share variants in genes in specific networks and pathways sufficient to give rise to a shared phenotype. We combined high throughput sequencing with advanced bioinformatic approaches to identify subgroups of patients with shared networks and pathways associated with preterm birth (PTB). We previously identified genes, gene sets and haplotype blocks that were highly associated with preterm birth. We performed targeted sequencing on these genes and genomic regions on highly phenotyped patients with 2 or 3 generations of preterm birth, and term controls with no family history of preterm birth. We performed a genotype test for differential abundance of variants between cases and controls. We used the genotype association statistics for ranking purposes in order to analyze the data using a multi-sample, protein-protein interaction (PPI) tool to identify significant clusters of patients associated with preterm birth. We identified shared interaction networks of proteins among 45 preterm cases in two statistically significant clusters, *p*<0.001. We also found two small control-dominated clusters. For replication, we compared our data to an independent, large birth cohort. Sequence data on 60 cases and 321 controls identified 34 preterm cases with shared networks of proteins distributed in two significant clusters. Analysis of the layered PPI networks of these clusters showed significant similarity scores between the clusters from the two independent cohorts of patients.

Canonical pathway analysis of the unique genes defining these clusters demonstrated enrichment in inflammatory signaling pathways, the glucocorticoid receptor, the insulin receptor, EGF and B-cell signaling, These results provide insights into the genetics of PTB and support a genetic architecture defined by subgroups of patients that *Share* variants in genes in specific networks and pathways which are sufficient to give rise to the disease phenotype.

**Author Summary:** The genetic architecture of complex diseases is reflected in subgroups of patients with variants in genes in specific networks and pathways. There are likely multiple networks that give rise to similar phenotypes. Preterm birth is an important complex genetic disease. We combined high throughput sequencing with advanced bioinformatic approaches to identify subgroups of patients with shared networks and pathways associated with preterm birth (PTB). We sequenced patients with 2 or 3 generations of preterm birth, and term controls with no family history of preterm birth. We used a novel protein-protein interaction network analysis to identify clusters of patients with shared networks in pathways for this important clinical problem. We identified shared interaction networks two significant clusters. We replicated these data, finding similar clusters, in an independent, large birth cohort.

## Introduction

Genome-wide association studies (GWAS) are a contemporary approach to the investigation of complex diseases that have made possible discovery of insights not previously recognized(1–5). However, GWAS have failed to demonstrate the “missing heritability” in many common diseases(6–10). The computational approaches underlying GWAS reflect the “common disease-common variant hypothesis,” that complex disease architecture is due to additive genetic effects of variants in individual genes. However, the genetics of complex diseases suggests that is unlikely. The more likely genetic architecture is that subgroups of patients share variants in genes in specific networks and pathways which are sufficient to give rise to a shared phenotype. It is also likely that variants in genes in different networks and pathways express similar phenotypes and define different subgroups of patients.

Preterm birth is an important, complex genetic disorder affecting up to 10% of pregnant women11. We previously used a bioinformatic approach to curate published literature and publicly available, high-throughput databases to develop a validated collection of genes with an *a priori* connection to preterm birth(11). We used gene set enrichment analysis (GSEA) with this refined gene set to analyze a large genome wide association study to identify biological networks and pathways associated with preterm birth(12). In order to identify the variants underlying these associations, we targeted the exons, flanking sequence and splice sites of the 329 genes and haplotype blocks that we showed were associated with preterm birth(12).Further, in order to leverage the likelihood of genetic discovery, we exploited an “extreme phenotype” of preterm birth by concentrating our enrollment on patients with a family history of preterm birth. We compared variants identified in women with 2–3 generations of preterm birth with term controls without history of preterm birth. We then used *Proteinarium*, a multi-sample, protein-protein interaction analysis (PPI) tool, to identify clusters of patients with shared PPI networks associated with preterm birth(13).

## Results

The clinical characteristics of the patients and their distribution by race/ethnicity are shown in Table 1.

**Table 1.** Clinical characteristics of patients

|  | **Cases N = 128 (%)** | **Controls N = 68 (%)** | **p-value** |
| --- | --- | --- | --- |
| Maternal Age | 27 ± 6 | 27 ± 7 | N.S |
| Gravida | 3 ± 2 | 2 ± 1 | N.S |
| Gestational Age wk | 31 ± 3 | 40 ± 1 | 0.001 |
| Birth Weight gm | 1724 ± 607 | 3451 ± 371 | 0.001 |
| African American | 15 (12) | 8 (13) | N.S. |
| Asian | 4 (3) | 3 (5) | N.S |
| Caucasian (non-Hispanic) | 75 (59) | 37 (60) | N.S |
| Hispanic or Latino | 30 (23) | 14 (23) | N.S |
| Native American | 1 (1) | 0 (0) | N.S |
| Other | 3 (2) | 0 (0) | N.S |

The only significant difference between the groups was in gestational age and birth weight. As described in the Methods, in order to leverage genetic discovery, the patients were carefully phenotyped with respect to history of preterm birth. The distribution of family history of preterm birth among the enrolled patients is shown in Table 2. Of the enrolled patients, 84 had a multi-generational history of preterm birth, 6 had an intra-generational history of preterm birth, 32 were first generation with multiple preterm births and 68 control patients had no family history of preterm births.

**Table 2.**
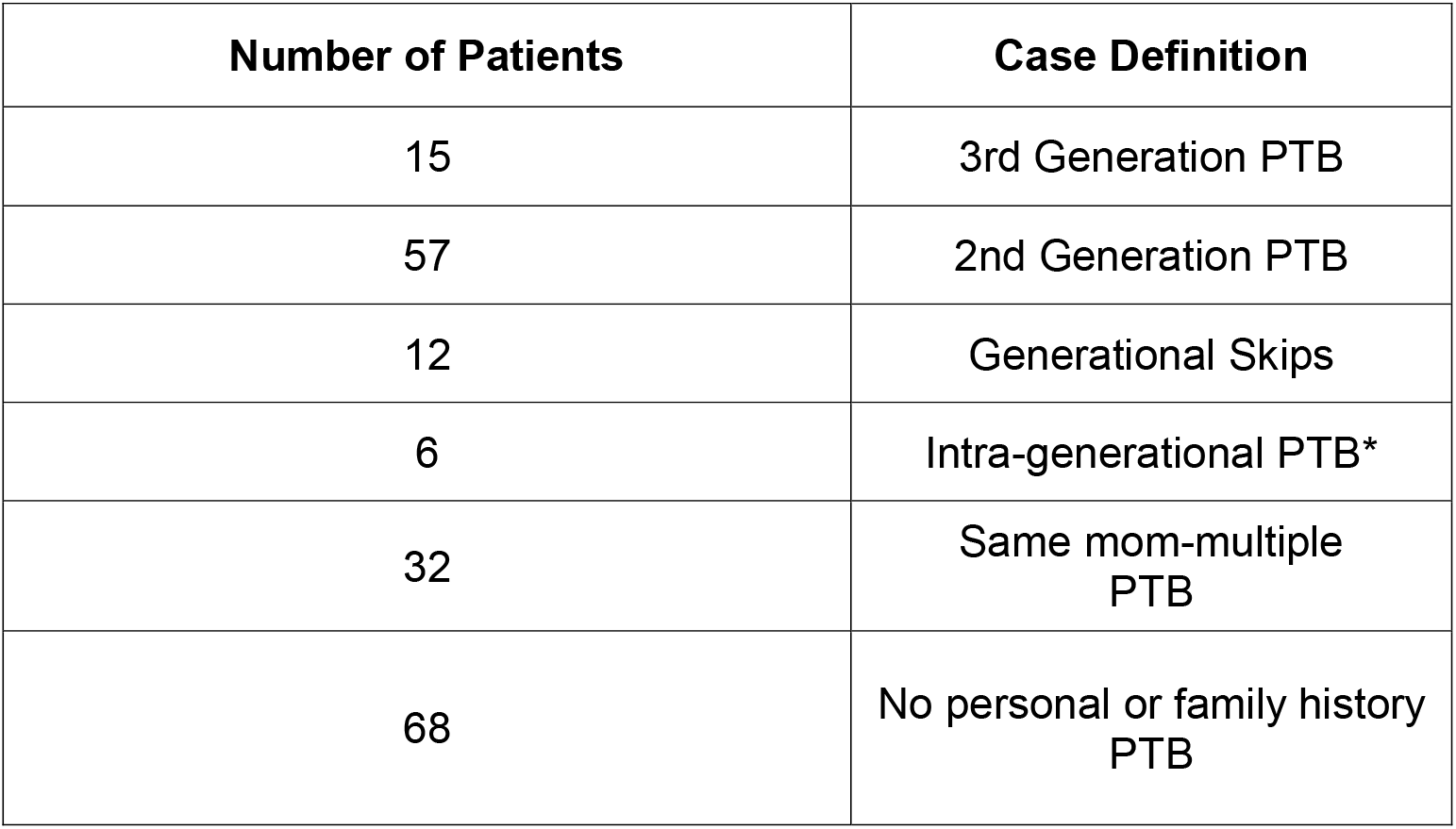
Family History of Preterm Birth among Enrolled Patients

| Number of Patients | Case Definition |
| --- | --- |
| 15 | 3rd Generation PTB |
| 57 | 2nd Generation PTB |
| 12 | Generational Skips |
| 6 | Intra-generational PTB* |
| 32 | Same mom-multiple PTB |
| 68 | No personal or family history PTB |

We identified a total of almost 140,000 variants, the bulk of which were in intronic regions captured from the haplotype blocks previously identified(12). We restricted our subsequent analyses to variants in regions with greater than 10-fold coverage which resulted in 39,472 variants. There were also almost 7000 exonic variants and several splice variants. After application of the initial filters for coverage and variant pathogenicity, there were a total of 264 variants (Supplemental Table 1). Of these, there were 9 variants that were nominally associated with preterm birth. All were non-synonymous, exonic variants (Table 3). A single SNV in the AOAH gene was more abundant in the cases, whereas the remaining eight variants were only present in the controls. Nonetheless, none met significance after correction for multiple comparison testing. None of the splice variants passed genotype testing for differential abundance between preterm cases and controls.

**Table 3.** Nominally significant genes from univariate analysis. Nonsynonymous SNV (nSNV)

| Chr | Gene | pos | Exonic<br>Function | Polyphen2_<br>HDIV_score | CADD_<br>phred | SIFT_<br>score | ExAC_<br>ALL |
| --- | --- | --- | --- | --- | --- | --- | --- |
| 1 | *COL16A1* | 32164127 | nSNV | 1 | 14 | 0 | 0.002 |
| 7 | *AOAH* | 36656035 | n SNV | 1 | 21 | 0.01 | 0.1172 |
| 9 | *QRFP* | 133769023 | n SNV | 0.999 | 18 | 0 | 0.0043 |
| 10 | *SORBS1* | 97144031 | n SNV | 1 | 17 | 0.01 | 0.0007 |
| 11 | *ATM* | 108098576 | n SNV | 0.98 | 18 | 0 | 0.0074 |
| 12 | *TBX5* | 114837349 | n SNV | 1 | 23 | 0 | 0.0034 |
| 16 | *CHST4* | 71571658 | n SNV | 0.999 | 17 | 0.02 | 0.0033 |
| 19 | *MYH14* | 50747534 | n SNV | 1 | 15 | 0 | 0.0029 |
| 20 | *TCFL5* | 61485507 | n SNV | 1 | 16 | 0 | 0.0028 |

For replication of these univariate data, we compared our results to a cohort of patients recruited at the INOVA Translational Medicine Institute, Falls Church VA(26, 27). They enrolled 816 families who underwent 60X whole genome sequencing. From these families, there were 60 cases and 321 controls that met our strict phenotypic criteria (singleton pregnancy, less than 34 weeks gestation, no history of preeclampsia or drug use). INOVA provided the variant data for the genes and haplotype block intervals described above. With similar functional filters, among the 264 variants identified from our cohort, 165 (63%) were also identified in the INOVA sequence data).

Among our cases with a family history of preterm birth, we found that each patient had an average of 163 variants that passed the coverage and Eigenstrat genotype testing above. In order to identify clusters of patients with shared networks associated with preterm birth, the top 30 genes based on the most significant variants (ranked by genotype p value) for each patient were used as the seed genes for input into *Proteinarium*. The resulting dendrogram is shown in Fig. 1. For ease of visualization the dendrogram has been circularized and the significant clusters have been highlighted in colors. The inset in Fig. 1 shows the distribution of cases and controls that belonged to each cluster. There were four significant clusters identified at a Fisher exact test with *p*< 0.001. Out of the 190 patients sequenced, a total of 66 subjects were assigned to statistically significant clusters. The two largest significant clusters (A and B) had significantly more cases than controls, encompassing 45 of the 128 cases. There were also two small control-dominated clusters. The layered networks for the case-dominated clusters A and B are shown in Fig.s 2a and 2b, respectively. The unique genes associated with these clusters are highlighted in light blue. There were 9 unique genes in cluster A and likewise 6 genes unique to cluster B (Table 4). All of the genes from the layered network graph of the two case dominated clusters and group membership of each gene are shown in Supplemental Table 2.

**Figure 1.**
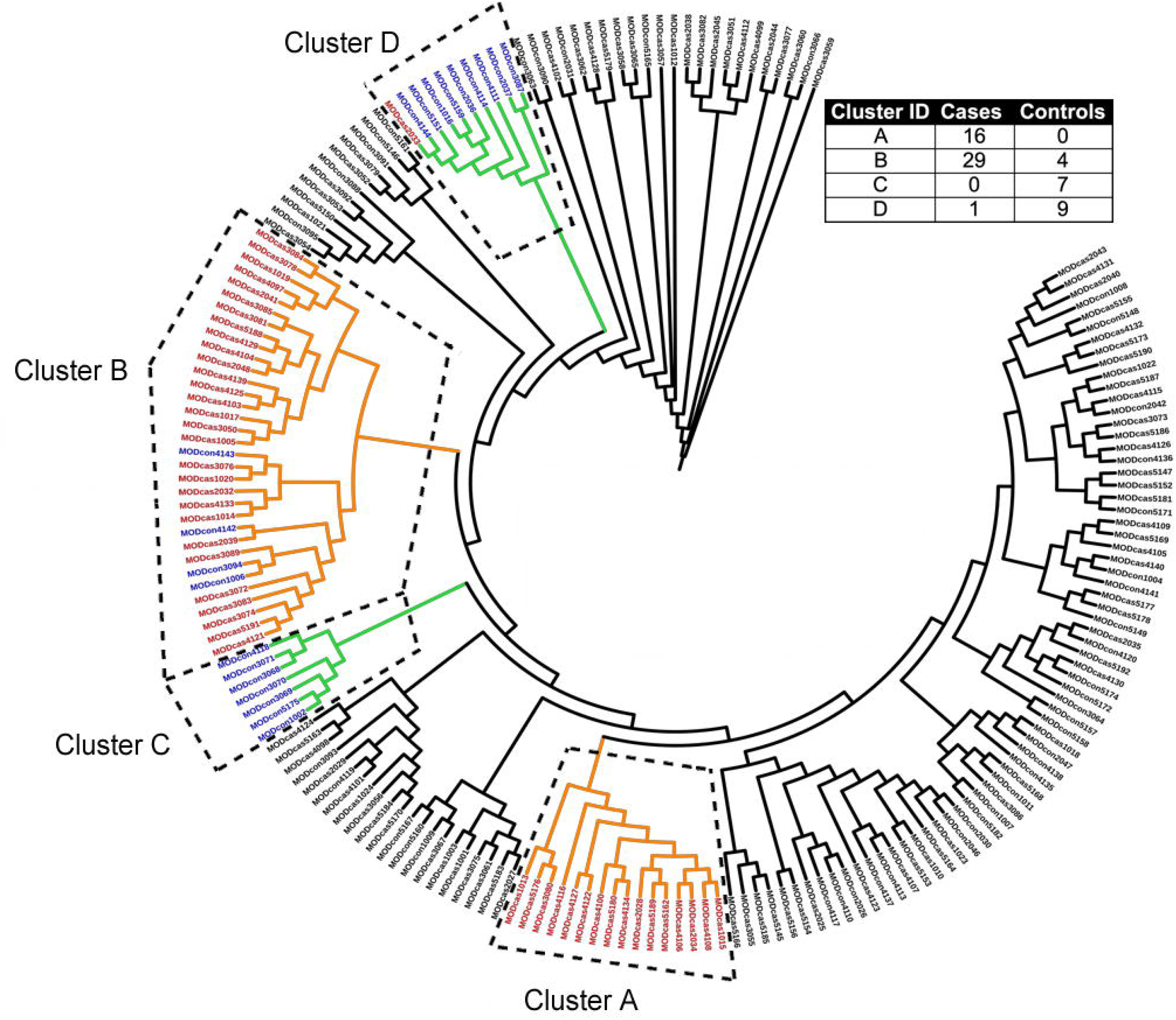
Dendrogram showing significant clusters of patients (colored). Inset: distribution of cases and controls in each of the clusters.

**Figure 2.**
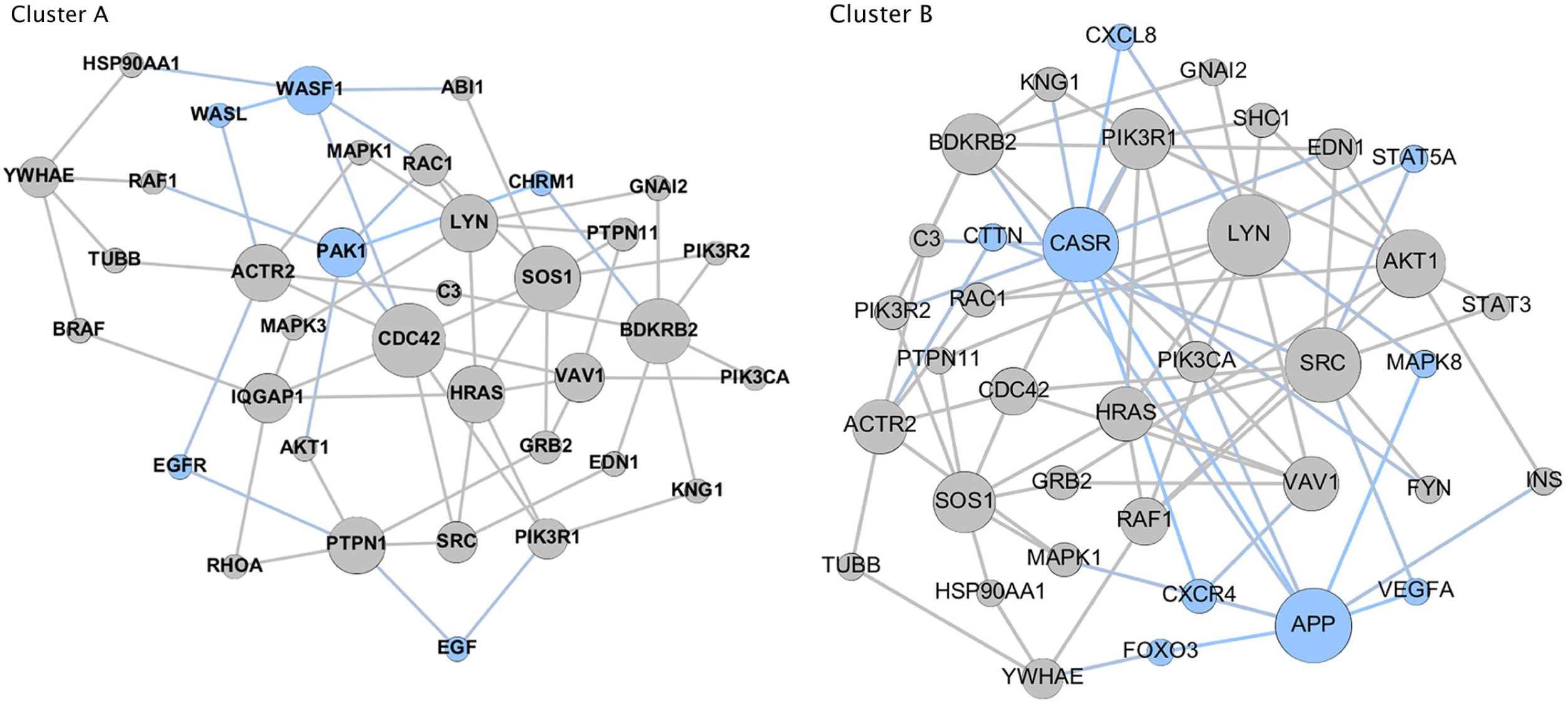
Layered network graphs for the case dominated clusters A & B fromFigure 1. The unique genes associated with each cluster are highlighted in light blue.

**Table 4.** Unique Genes from case dominated preterm birth clusters

| **Genes\*** | **Cluster** | **Sequenced** | **Imputed** |
| --- | --- | --- | --- |
| *APP* | A | Yes | No |
| *CASR* | A | Yes | No |
| *CTTN* | A | No | Yes |
| *CXCL8 (IL-8)* | A | No | Yes |
| *CXCR4* | A | Yes | Yes |
| *FOXO3* | A | Yes | No |
| *MAPK8* | A | No | Yes |
| *STAT5A* | A | No | Yes |
| *FOXO3* | A | Yes | No |
| *CHRM1* | B | No | Yes |
| *EGF* | B | No | Yes |
| *EGFR* | B | Yes | No |
| *PAK1* | B | Yes | No |
| *WASF 1* | B | Yes | No |
| *WASL* | B | No | Yes |

\*Alphabetically ordered for each cluster

For replication of this network analysis, a similar approach using the filters described above to identify variants in individual patients was applied to the INOVA cohort. The top 30 seed genes for each subject were used for input into *Proteinarium*. There were four significant case-dominated clusters identified encompassing 40 of the 60 cases. The layered networks for these four clusters are shown in Fig. 3. The unique genes associated with these clusters are highlighted in light blue. The gene lists for the layered networks for the case dominated clusters from the preterm birth cohort and the replications cohort and group membership is shown in Supplemental Table 2.

**Figure 3.**
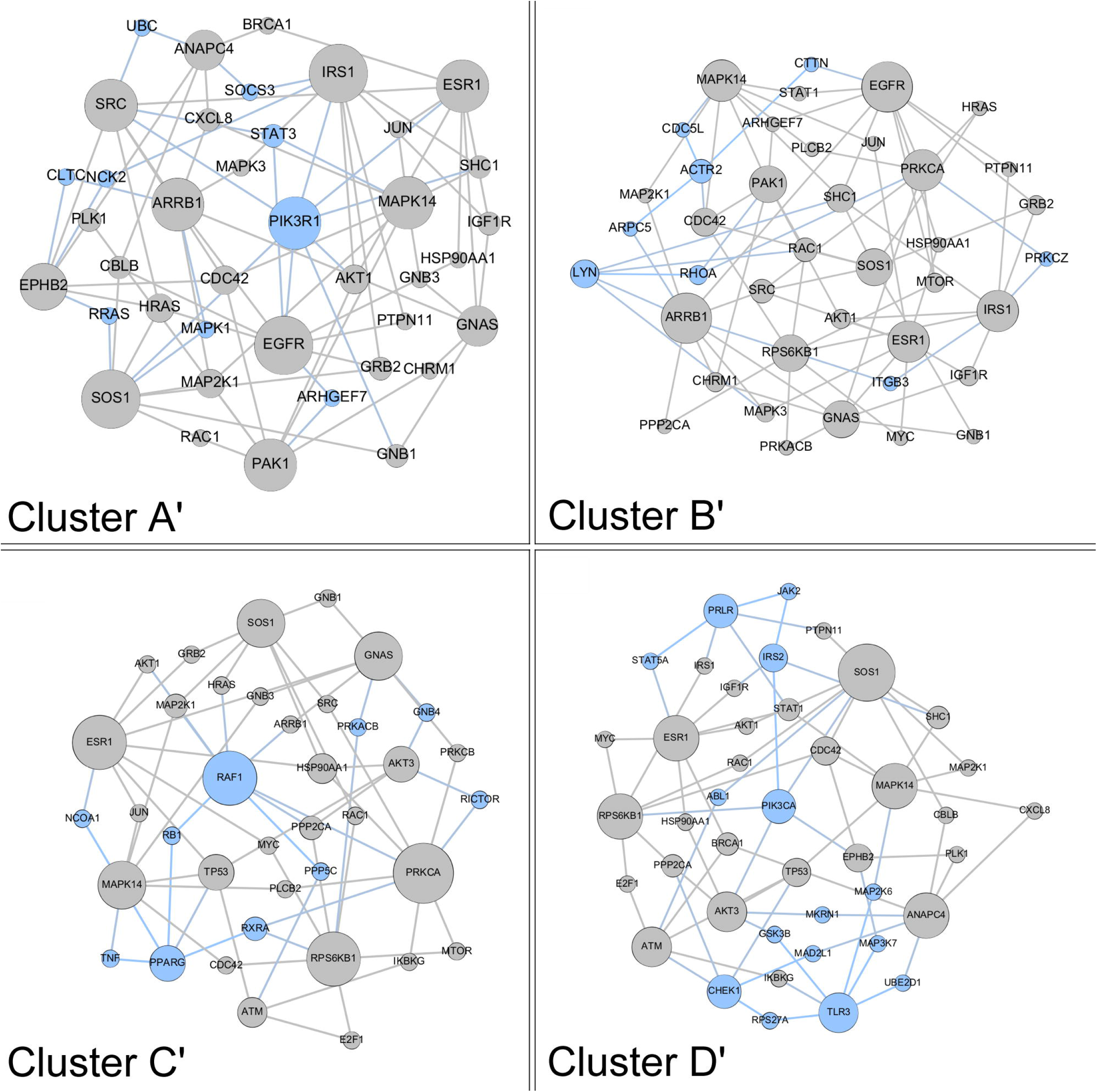
Layered network graphs for the INOVA replication cohort showing significant clusters A’, B’, C’, D’. Unique genes to each cluster are shown in light blue.

We used separation testing to compare the networks identified by *Proteinarium*. The two case dominated clusters from our preterm birth cohort (clusters A and B) showed overlap with each other within the interactome. This was reflected by negative separation values (separation = –0.285) when compared to the control dominated clusters that showed higher separation values (ranging from –0.037 to 0.11).

We also compared the separation of the case dominated clusters from our preterm birth cohort to the case dominated clusters in the INOVA replication cohort. The results are shown in Table 5. Case clusters B’ and D’ from the replication cohort showed the greatest similarity when compared to our case clusters B and A, respectively, indicating significant overlap within the interactome.

**Table 5.** Separation scores for comparison of case dominated clusters from the preterm birth cohort and the replication cohort.

|  | Replication Cohort |  |  |  |  |
| --- | --- | --- | --- | --- | --- |
|  |  | A' | B' | C' | D' |
| Preterm Birth Cohort | A | -0.224 | 0.010 | -0.193 | **-0.330** |
|  | B | 0.114 | **-0.491** | -0.123 | -0.239 |

In order to identify the pathways modulated by the genes defining the networks within significant clusters of cases, we carried out comparative pathway analysis for the genes defining each cluster using Ingenuity Pathway Analysis^Tm^. For this analysis we included the genes unique to each of the layered network graphs of the two case dominated clusters(clusters A and B) in the preterm birth cohort and the two case dominated clusters in the replication cohort that had the smallest separation with those clusters (clusters B’ and D’). To avoid false positive inferences, we used stringent criteria with *p* values ranging from 10^^6^ to 10^^8^. The pathway similarity is illustrated in the heatmap shown in Fig. 4. Canonical pathway analysis of the unique genes defining these clusters demonstrated enrichment in numerous signaling pathways. This includes signaling by IL6, IL-7, IL-15, IL-8, JAK/Cytokine signaling, toll-like receptors, the glucocorticoid receptor, the insulin receptor, EGF and B-cell signaling.

**Figure 4.**
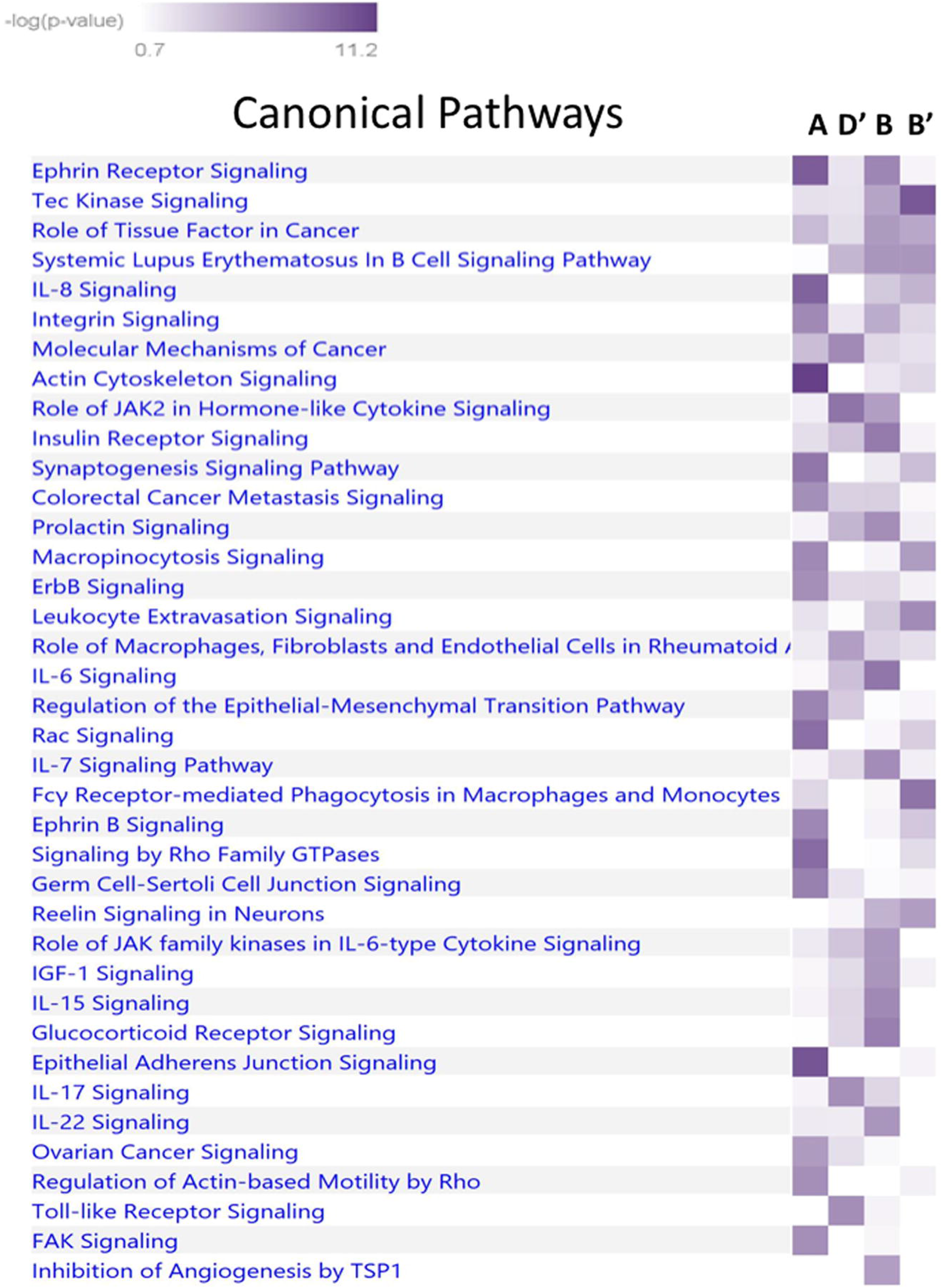
Comparative network analysis from Ingenuity Pathway Analysis. Comparison of case dominated clusters A, D,’ B, B’. All pathways significant *p* 10^^6^ to 10^^8^.

We also sought to determine if there were underlying phenotypic differences between the patients in the different clusters we identified using *Proteinarium*. The clinical phenotype of the case patients in Cluster A, were compared to the characteristics of the remaining patients not included in Cluster A or Cluster B. An analogous analysis was performed for Cluster B. In addition, the characteristics of patients in Clusters A and B were compared to each other. The distribution of clinical characteristics for these comparisons is shown in Table 6. Comparing the cases from Cluster A to the subjects in neither of the two clusters, there was a significant difference in the distribution of maternal racial background and in the generational history of preterm birth (p<0.05). For Cluster B compared to the subjects in neither of the two clusters, there was a significant difference in the proportion with chorioamnionitis (p<0.05). Comparison of the patients in Cluster A and Cluster B, showed a significant difference in income and the distribution of maternal racial background (p<.05). Nonetheless, the majority of these differences in clinical characteristics were very modest.

**Table 6.**
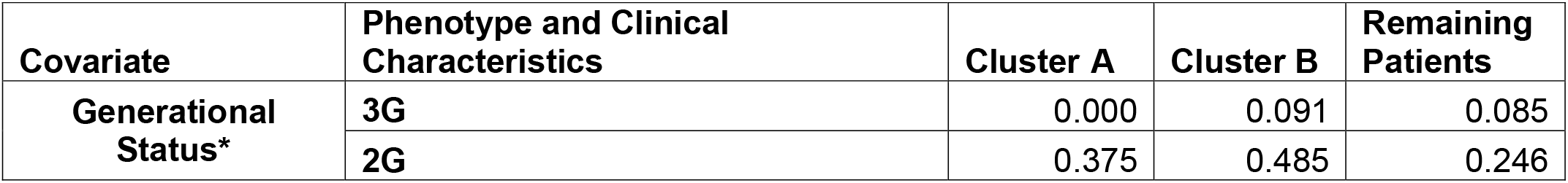
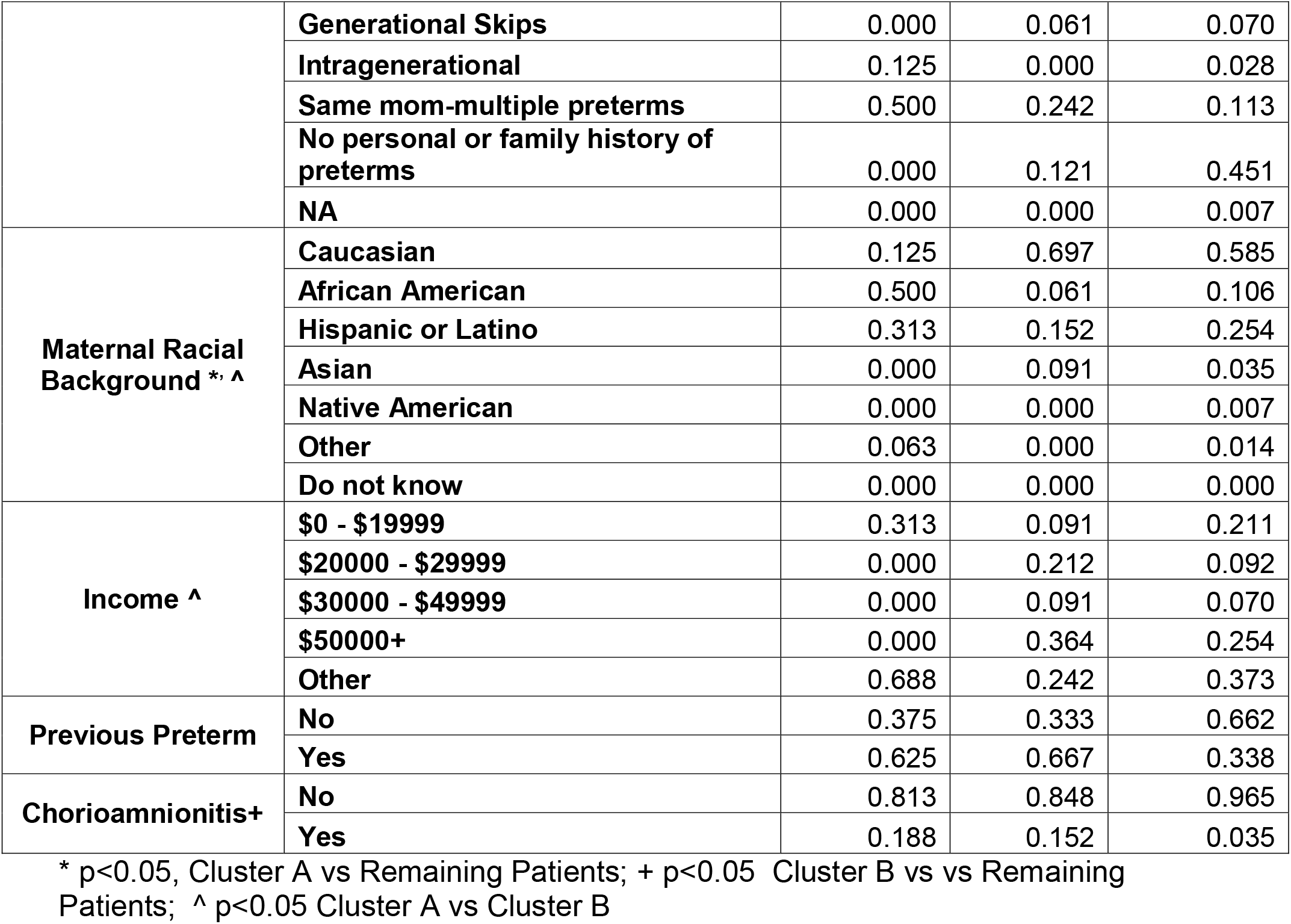
Proportion of patient clinical characteristics within clusters

## Discussion

We performed targeted sequencing of gene sets and haplotype blocks that are highly associated with preterm birth on carefully phenotyped patients. We enrolled women with idiopathic, singleton births <34 weeks gestation, the majority of whom had multiple generations of preterm birth. We compared them to term controls with no family history of preterm birth. We used *Proteinarium*, a multi-sample, protein-protein interaction tool, to identify clusters of patients with shared protein-protein interaction networks associated with preterm birth(13). Using seed genes from each patient, *Proteinarium* mapped the input genes onto the STRING PPI interactome to build individual networks. The similarities between all subjects’ PPI Networks were used as the distance metric for clustering samples. We identified two significant clusters with a predominance of preterm birth patients encompassing 45 out of the 128 women with a multi-generation history of preterm birth. We also found two small control-dominated clusters. For replication, we compared our data to a large birth cohort collected at INOVA Health (27). Sequence data analyzed from INOVA’s 60 cases and 321 controls identified 40 preterm cases in four significant clusters. Separation analysis of the layered PPI networks of the significant clusters from our preterm birth cohort and the replication cohort demonstrated overlap of these clusters within the interactome. Canonical pathway analysis of the unique genes defining these clusters demonstrated enrichment in inflammatory signaling by *IL-6, IL-7*,*IL-15, IL-8*, JAK/Cytokine signaling, toll-like receptors, the glucocorticoid receptor, the insulin receptor, EGF and B-cell signaling, *p∼* 10^−6^ to 10^−8^.

The individual, unique genes from the two case-dominated preterm birth clusters are shown in Table 4. Several of the genes unique to Cluster A have been association with inflammation and immune responses. *CXCR4* is among the gene sets and pathways upregulated in whole blood from women with spontaneous preterm birth when compared to patients delivering at term(28). Moreover, *CXCR4* is located in genomic regions with large ROH that we have shown to be in greater abundance in women delivering preterm than full term(29).*CXCR4* has also been evolutionarily linked to preterm birth(30). *CXCL8* (IL-8) is a monocyte macrophage chemoattractant. It has been widely studied in labor and shown to be expressed in multiple gestational tissues including myometrium, cervix and decidua(31). *CXCL8* is upregulated in chorio-decidua samples collected from preterm labor patients when compared to patients at term and in labor(31). This is consistent with the increase in inflammatory cells in the decidua during labor. The gene *FOXO3* codes for the forkhead box 03 transcription factor that regulates inflammation in non-gestational tissues. It has also been shown to be expressed in human myometrial tissue(32). Further, higher *FOXO3* gene and protein expression have been demonstrated in myometrium from women in labor compared to non-laboring samples. In isolated cells, *FOXO3* silencing was associated with a significant decrease in IL-1 induced I-L6 and IL-8 expression and cyclooxygenase 2 production. Overexpression of *FOXO3*, increased cytokine expression, prostaglandin production and *MMP9* expression are all observed in myometrial cells following administration of IL-1B. Thus, *FOXO3* may be implicated in the pathways regulating labor and it may be a potential target for prevention of preterm birth(32).The *STAT5A* gene is present in a network of cell-mediated immune responses, cellular movement and hematologic system development that was identified in a genome-wide association study looking at single nucleotide polymorphisms (SNP) in peripheral blood of patients who are in preterm labor compared to full term labor(33). Most importantly, serial sampling and transcriptional profiling of circulating immune cells during pregnancy has been carried out to identify patterns of gene signatures across pregnancy that are associated with gestational age and thus define a “gestational clock” (34). These studies revealed an important role for IL-2 dependent *STAT5A* signaling in modulating T-cell function during pregnancy.Baseline gene expression was compared with expression following activation with receptor-specific ligands including interferon and a cocktail of interleukins. The endogenous *STAT5A* signal in naïve cells and the phospho-*STAT5A* response to interleukins and neutrophils were among the strongest features correlated to immunological adaptations to pregnancy and association with gestational age(34). The Comparative Toxicogenomics Database (CTD) contains more than 5,000 curated and inferred gene–disease associations (including preterm birth) extracted from the published literature by formal curation(35). *STAT5A* is also among the CTD gene disease phenotypic associations with preterm birth. MAPK8 is also among the CTD gene disease associations associated with preterm birth(35). Lastly, several genes in Cluster A are involved in cellular migration and invasion, which may be important in the onset of labor.*CTTN* is the cortactin gene. It functions as a key regulator of actin cytoskeleton and has roles in actin-based cellular processes including cell migration and invasion. Polymorphisms in the *VEGFA* gene in a discrete haplotype block have been shown to be associated with preterm birth(33). This may play an important role in angiogenesis during placentation. Nonetheless, the latter study was of modest size and the findings just reached nominal significance.

In support of the network analysis, many of the genes are involved in uterine contractility, signal transduction and cell-cell signaling. Moreover, there is substantial literature based evidence that 4 out of the 6 unique genes in Cluster B are involved in uterine contractility. Signaling via *EGF* and the EGF receptor in human amnion cells regulates their proliferation and increases calcium mobilization and PGE2 production which may also have significant effects on uterine contractility(36). *PAK1*, one of the genes identified in the Database for Preterm Birth(11), encodes a member of the serine threonine P21 activating kinases. *PAK1* is only present in pregnant myometrial tissue. PAKs have shown to regulate uterine contractility(37). We have also previously reported that *PAK1* is located in a genomic region with runs of homozygosity (ROH) which are significantly more abundant in mothers delivering preterm than term(29). Activation of the *CHRM1* receptor by acetylcholine increases uterine contractility(38). Of functional significance, *CHRM1* has been shown to be down-regulated in preterm human myometrium compared to patients at term and not-in-labor(39). *WASF1* is an A-kinase anchoring protein. It has been implicated in preterm labor by the Ontario Birth Study where it was shown to be differentially expressed in patients undergoing preterm labor(40). It was further shown to be responsive to glucocorticoids in another study of peripheral blood mononuclear cells from patients delivering preterm(40).

There have been many attempts in the past to identify genetic contributions to the risk of preterm birth(17, 41–46). Polymorphic changes in the protein coding regions, regulatory and intronic sequences of specific genes have been described. In most studies, candidate genes or proteins involved in inflammatory reactivity or uterine contractility have been investigated(17, 41–58). The results suggest that alteration in the expression of these proteins interacts with infection and/or other environmental influences associated with preterm birth. The results however, do not provide insight into the causes of prematurity in the absence of inflammation or infection. Moreover, while interventions directed at infection or inflammation have been successful in experimental models they have largely not been successful in treatment or prevention of preterm birth in humans(54).

More recently, investigations using high throughput and multi-omic techniques have been undertaken. Sakabe et al compared transcriptome and regulatory maps of decidua-derived stromal cells to a genome-wide association study of gestational duration(59). Using a combination of techniques, including ATAC-seq, Hi-C and ChIP-seq, they identified the chromatin landscapes in decidua-derived stromal cells. These were then compared to the heritability of annotations in the GWAS of pregnancy-related traits. They showed the heritability of gestational-duration was enriched for functional annotations in decidual stromal cells(59).Volozanoka recently carried out targeted sequencing of genes shown to be related to cervical insufficiency following a systematic literature analysis similar to that underlying this study(11, 60). They identified 12 genes that were normally linked to cervical insufficiency. However, this was a modest study involving only 21 patients and there were no overlaps with our gene set.Zhou et al recently analyzed publically available gene expression data sets derived from maternal blood in the second and third trimesters of women with spontaneous preterm birth and term birth(61). Expression of a single gene, *EBF1*, was associated with preterm birth(62). In a large genome-wide association study of over 1300 cases of spontaneous preterm birth in comparison to 12,000 ancestry-matched controls, they identified only two intergenic loci associated with preterm birth. The authors concluded that the genetic contributions to preterm birth are unlikely due to single common genetic variants but could be explained by interactions of multiple variants or environmental influences(62). Meta-analysis of maternal and fetal transcriptomic data from genomic databases for studies related to preterm birth identified genes differentially expressed in spontaneous preterm birth relative to term^63^. Ontogeny analysis demonstrated that maternal changes were enriched in immune-related pathways, upregulation of innate immunity and downregulation of adaptive immunity(63). By comparison, analysis of the transcription profile in cord blood showed a downregulation of innate immune findings.Nonetheless, the results demonstrate a significant relationship of immune functions in the pathogenesis of preterm birth. Multi-omic analysis of preterm birth from the parent study of our INOVA replication samples has been reported(27). It was a large study integrating whole genome sequencing, RNA sequencing and DNA methylation data for 270 preterm births and 520 control families. In their univariate analysis there were no variants that reached genome-wide significance. However, there were groups of genes associated with preterm birth in subsets of patients that were identified in secondary analyses(27). Integration of the three data sources (WGS, RNAseq, DNAm) identified a set of 72 candidate biomarker genes for very early preterm birth (VEPTB) and genes associated with PTB. *RAB31* and *RBPJ* were identified by all three data types in preterm birth patients. Additionally, pathways associated with VEPTB included the EGFR and prolactin signaling pathways, inflammation- and immunity-related pathways, chemokine signaling, IFN–γ signaling, and Notch1 signaling(27).

Our study has many strengths which contributed to successful identification of genetic variants in genes in networks associated with preterm birth. First, we used a very carefully phenotyped cohort of women with a strong family history of preterm birth. Our control cases were as carefully ascertained to have no family history of preterm birth. Second, we signaling, and Notch1 signaling(27). carried out targeted sequencing on genes with a demonstrated role in preterm birth. Third, we employed a novel analysis of protein-protein interactions to identify clusters of patients with shared PPI networks associated with preterm birth. Our study also has limitations and areas that deserve consideration. The significant clusters of preterm birth cases included unique genes that were both in our targeted sequencing as well as imputed via network analysis. Even though not included in our original sequencing, the fact that the unique imputed genes had a strong association with preterm birth, uterine contractility and immune responses is noteworthy. This was a modest sample size. We identified significant clusters of patients with networks and pathways associated with preterm birth but those findings were restricted to 45 out of the 128 cases. We did not anticipate being able to assign each of the cases to a significant cluster. We believe that the targeted sequencing contributed to our successful discovery even though all cases were not assigned to significant clusters. Nonetheless, the fact that we were able to identify significant case dominated clusters in our preterm birth cohort at all and that we were able to demonstrate similarity to clusters of patients with shared networks in the replication cohort lends validity to our hypothesis on the genetic architecture of this complex disease and the genetic leverage provided by the family history of preterm birth. The fact that this was not a whole exome study likely contributed to the number of cases we were able to assign to significant clusters. Futures studies employing similar techniques but with whole exome sequencing are likely to expand the number of case clusters that we will be able to identify using this approach. It is beyond the scope of this report to thoroughly discuss the control dominated clusters. Nonetheless, several elements deserve mention. We interpret the networks and pathways that are shared between control patients to represent protective genes or genes that confer resiliency against preterm birth. It is notable that we were able to identify significant clusters in the controls from our preterm birth cohort but none were identified in the replication cohort. We attribute this to the careful phenotyping that was used to enroll patients in the primary study.

In summary, we used a novel multi-sample, protein-protein interaction tool, to identify clusters of patients with shared protein-protein interaction networks associated with preterm birth. We showed similarity between these networks and results from an independent replication cohort. Our results provide insights into the genetics of PTB and support a genetic architecture defined by subgroups of patients that *share* variants in genes in specific networks and pathways which are sufficient to give rise to the disease phenotype.

## Material and Methods

### Patient Identification and Enrollment

Large epidemiological studies drawn from population based analyses support a predominantly maternal origin for the genetic contribution(s) to risk of preterm birth, with little contribution by paternal or fetal genetic factors(14–17). We therefore concentrated our efforts on identification of maternal genetic variants. Women & Infants Hospital of Rhode Island is the sole provider of high-risk perinatal services in Rhode Island, northeastern Connecticut and southeastern Massachusetts. We used this *population-based* service to enroll patients with a prior history of preterm birth. The study was approved by the Institutional Review Board, 08-0117. An informatically driven retrieval from our electronic medical record gave us a daily report on all preterm births. A clinical research assistant, formally trained in genetic interviews, reviewed the records of all patients delivering < 34 weeks. Following informed consent, women underwent an interview focused on family history of preterm birth. We asked explicit questions about preterm birth in mother, grandmother, her first order relatives and also paternal relatives. Careful clinical history with an emphasis on additional risk factors for prematurity including medical illnesses, drug use, psychiatric disorders and employment history was recorded on all patients. We excluded patients delivered prematurely for considerations related to preeclampsia, drug use, diabetes or multiple gestation. Controls were patients who delivered ≥ 37 weeks gestation in whom the same, formal genetic history revealed no history of preterm birth on either maternal or paternal side of the pedigree. All of the patients’ identifying data was coded and redacted for the purposes of data analysis. 190 patients were enrolled for targeted sequencing. Samples were taken from 128 women with multiple generations of preterm birth, and 68 race, ethnicity matched control women at term. Residual maternal whole blood was obtained for extraction of genomic DNA. The samples were stored continuously at –80°C until processing.

### Sample preparation

We targeted the 329 genes and 132 haplotype blocks for sequencing that are highly associated with preterm birth(12). Genomic DNA from maternal whole blood was extracted using QIAamp DSP DNA blood mini kit from Qiagen following the manufacture’s protocol.Samples were quantified using Qubit technology (Life Technologies, Carlsbad, CA, USA) and sequencing libraries were constructed from 2 μg each of case/control DNA. Library preparation was performed using Illumina TruSeq DNA LT Sample prep Kit (Illumina, San Diego, CA, USA), with enzymatic fragmentation using dsDNA Fragmentase (NEB), followed by indexing and clean-up. DNA capture was performed using custom capture probes from SeqCap EZ choice kit(Roche NimbleGen).

### Targeted sequencing

The library was sequenced on an Illumina HiSeq 2500 using 100 bp paired-end protocols. Following sequencing, the multiplex indices were used to bin the samples for each patient and QC sequence data was recorded. High quality sequence data from well-balanced pools was observed. There was an average of 25 million reads from each patient, with an ≥ average Q30 of 91%. Reads were then mapped to the to the human reference sequence(Hg19) with BWA(18),sorted and indexed with SAMtools(19).

### Sequence data, variant calling and genotype testing

Variants were flagged as low quality and filtered using established metrics: if three or more variants were detected within 10bp; if four or more alignments mapped to different locations equally well; if coverage was less than ten reads; if quality score < 30; if low quality for a particular sequence depth (variant confidence/unfiltered depth < 1.5); and if strand bias was observed (Phred-scaled p-values using Fisher’s Exact Test > 200). A variant identified by any one of these filters was labeled “low quality” and not considered for further analysis. For variant discovery we used the Gene Analysis Tool Kit (GATK) version 3.2 to analyze the sequence reads(20). Duplicate reads were marked and removed using Picard Tools version 1.77.Haplotype caller was applied for variant detection(21). Twenty-five base pairs upstream and downstream of each exon were included in the design of capture probes and in variant detection. Variants were annotated using ANNOVAR for pathogenicity prediction scores. We used Eigenstrat to control for population stratification during genotype testing of differential abundance of variants in cases and controls(22). To investigate the frequency of potentially relevant single variants, we extracted variants with the following filters: coverage ≥10X, a Polyphen 2 HDIV prediction if a change is damaging (>=0.957), a SIFT score (<0.05), a CADD score > 10, and minor allele frequency (MAF) <0.05 from the Exome Aggregation Consortium (ExAC)(23), and significant difference by genotype testing^22^.

### Network Analysis

In order to identify patients with shared networks and pathways associated with preterm birth, we used *Proteinarium*^(13)^. This is a tool for analysis of protein-protein interactions that uses the String interactome database, Dijkstra’s algorithm and the Jaccard index to build a network similarity matrix of protein-protein interactions (PPI) between samples(13). The top 30 genes, based on the most significant variants (ranked by Eigenstrat genotype p value) for each patient, were used as the seed genes for input into *Proteinarium*. We selected *Proteinarium’s* user defined output minimum path length of 2, which includes pathways in which seed proteins are connected directly to each other and/or via a single intermediary protein. We refer to these intermediary connecting proteins as imputed proteins. The output of *Proteinarium* is a UPGMA generated dendrogram that shows clusters of patients with shared PPI networks, gene lists forming the networks and the group assignment for each patient. Statistical significance for each branch under the dendrogram is calculated by Fisher exact test comparing the probability of observing a cluster of that size relative to the total number of samples and group assignment.

### Network Separation Testing

Computational methods have been developed to identify disease-disease similarity by comparison of individual networks from the protein-protein interactome(24). This network-based approach compares the shortest distances between proteins *within* each disease or network to the shortest distances *between* the disease networks. This approach has been applied to other complex disease phenotypes(25). We computed the separation between our clusters using the seed genes identified for each of the patients. Using the union of the seed genes for patients within each of the clusters, we identified genes unique to each cluster to use as input for the separation analysis(24). A negative score indicates an overlap between networks within the interactome. The more negative the score the greater the similarity/overlap between two networks.

### Phenotypic Analysis

In order to identify significant differences in the clinical, phenotypic characteristics between the clusters identified by *Proteinarium*, we used a Bayesian generalized linear model implemented via the Arm package in R (Arm). The optimal model was determined using stepwise model selection with the MASS package in R (MASS).

## Data Availability

All relevant data referred to in the manuscript are avaialable.

http://ptbdb.cs.brown.edu/dbptb/

## Supplemental Material Legends

**Supplemental Table 1**. Functional annotation and variant numbers corresponding to pathogenicity and genotype filtering.

**Supplemental Table 2**. Genes and group membership of the genes from the layered network graph of the two case dominated preterm birth clusters (A and B) and the four case dominated clusters from the replication cohort (A’, B’, C, D’)

